# The Alzheimer’s disease Burden in China (ABC) study: protocol for a nationwide multicentre cross-sectional and prospective cohort study

**DOI:** 10.64898/2026.04.30.26352138

**Authors:** Yixuan Wang, Qi Qin, Kun Yang, Mingzhi Yu, Yao Yao, Chao Gong, Jing Guo, Li Yang, Yi Tang

**Author notes:** Corresponding author: Yi Tang, Department of Neurology & Innovation Center for Neurological Disorders, Xuanwu Hospital, Capital, Medical University, National Center for Neurological Disorders, Beijing Key Laboratory of Digital Medicine for Cognitive Disorders, 45 Changchun Street, Beijing 100053, China, Li Yang, Department of Health Policy and Management, Peking University School of Public Health, Beijing Institute for Health Development, Peking University, National Health Commission Key Laboratory of Health System Reform and Governance (Peking University), 38 Xueyuan Road, Beijing 100191, China. These authors have contributed equally to this work (Yixuan Wang, Qi Qin).

## Abstract

**Introduction:** Alzheimer’s disease (AD) is imposing an increasing public health and socioeconomic burden. In China, rapid population ageing is sharply increasing disease burden. Previous studies have shown that AD-related costs are mainly driven by long-term informal care. However, evidence in China remains limited by an incomplete cost framework, and insufficient consideration of caregivers’ burden and indirect costs. Notably, the *National Dementia Action Plan (2024–2030)*, issued by the Chinese government, marks a major shift to early detection and comprehensive care of AD, highlighting the urgent need for nationally representative economic evidence to support policy implementation. This study aims to evaluate the economic burden and quality of life of AD patients and their caregivers in mainland China, and is the first nationwide study to include individuals with amnestic mild cognitive impairment (aMCI), providing foundational data for future health technology assessment (HTA) of early AD interventions.

**Methods and analysis:** Baseline characteristics will be presented and compared using t-tests or chi-square tests. Economic burden will be estimated by calculating the per capita cost and weighted national total based on provincial numbers of AD patients. Indirect costs will be assessed using locally adapted replacement cost approach and forgone wages approach. The analysis will be stratified by disease severity and age. Future burden will be projected by linking data from China Statistical Yearbook 2025 and the United Nations World Population Prospects 2024. Unmet care needs, AD-related catastrophic health expenditure (CHE), and AD dependency ratio (ADDR) will also be assessed.

**Ethics and dissemination:** Ethics approval was obtained from the Ethics Committee of Xuanwu Hospital, Capital Medical University. The study has been registered at ClinicalTrials.gov and the Chinese Clinical Trial Registry (ChiCTR). The results from this study will be actively disseminated through research articles and conference presentations.

**Trial registration number:** **NCT05995418; ChiCTR2300074723**.

**Strengths and limitations of this study:**

- This study employs a two-stage probability sampling design across 602 cognitive centres in 31 provinces, ensuring strong national representativeness of the Chinese population.
- It is the first national health economics study to prospectively include patients with amnestic mild cognitive impairment (aMCI), addressing a critical evidence gap in the early stage of disease and providing data for future evaluation of the cost-effectiveness of early screening and interventions.
- The integration of clinical characteristics, economic burden, and quality of life scales provides a multidimensional framework for future policy evaluations and health technology assessment (HTA).
- The calculation of indirect costs relies heavily on caregivers’ self-reported data regarding care time and missed work, which may introduce recall bias.
- The use of a nested subsample for the 12-month follow-up may introduce loss-to-follow-up bias, although appropriate statistical weighting techniques will be applied to mitigate this.

## 1. Introduction

### Background

Alzheimer’s disease (AD) is the most common neurodegenerative disease. It causes progressive decline in cognitive function, ultimately leading to significant disability or death [1]. In China, the prevalence of AD and related dementias (ADRD) is increasing rapidly in the context of population ageing. As the disease progresses, patients require growing levels of care and assistance, resulting in escalating informal caregiving time and non-medical expenditures [2], [3]. Families often face difficult trade-offs between employment, caregiving responsibilities, and financial sustainability [4]. As a result, AD burden extends beyond clinical impairment to encompass substantial social and economic consequences, particularly for family caregivers [5].

Several health economic studies on AD have been conducted in China, among which the study by Jia et al. first highlighted the economic burden of AD in China, estimating direct medical, direct non-medical, and indirect costs [6]. This study laid a crucial foundation for domestic health economics research on AD. However, China’s population aging accelerated significantly over the past decade, economic development progressed steadily and healthcare systems and cognitive diagnosis capabilities improved continuously. The *National Dementia Action Plan (2024–2030)* issued by the Chinese government in 2024, marks a shift in China’s AD prevention and control policy. It has shifted from a single disease treatment to a full-course management including screening, diagnosis, treatment and follow-up [7].

Against this background, the “Alzheimer’s disease Burden in China” (ABC) Study is designed as a nationwide, multi-centre research integrating Hospital Information System data, structured questionnaire, and follow-up data. The ABC Study aims to cover multiple dimensions including clinical characteristics, economic burden, psychological scales, and policy impacts. This protocol describes the design, sampling strategy, variable framework, and planned analyses of the ABC Study, with the aim of providing a transparent methodological reference for AD health economics research in China. We aim to provide solid data support for future evaluation of intervention measures, design of long-term care insurance policies, and cross-disciplinary research.

### Current evidence gaps

Since the pivotal study by Professor Jia’s team in 2018, studies of economic burden of AD in China have entered a phase of rapid growth. More research has begun to explore the impact of disease staging on cost structures, regional resource allocation disparities, and the cost-effectiveness of new diagnostic technologies such as biomarkers. However, existing research exhibits several limitations that hinder the translation of study evidence into policy.

First, representative data across all provinces are still lacking, hindering equitable resource allocation and policy formulation between regions [8]. Second, insufficient stage-specific cost analysis fails to accurately reflect changes in burden structure across different disease stages [9]. Third, inadequate attention to early-onset patients and those with multiple comorbidities may lead to underestimation of indirect costs and family risks. More critically, existing studies often overemphasize economic expenditures while neglecting patients’ psychological well-being, quality-of-life changes, and caregivers’ emotional distress and life imbalances [10]. Without systematic assessment of these human-centered dimensions, estimated economic burdens fail to authentically reflect the disease’s comprehensive impact on families and society.

### Policy Urgency and Public Health Significance

In China, the *National Dementia Action Plan (2024–2030) has* set the goal of establishing a comprehensive, continuous prevention and control system for dementia by 2030. Therefore, policymakers require reliable, comprehensive, and regionally representative AD burden data support: How do we evaluate the incremental costs of population screening? How should Long-Term Care Insurance payment scope be tailored for individuals with cognitive impairment? How will the inclusion of novel AD drugs in the national medical insurance catalog impact total societal expenditure?

In response to this pressing need, the ABC Study emerges as a nationwide, multicenter investigation. This research will collect structured cost assessments, caregiver modules, and quality-of-life evaluations covering all disease stages. By combining cross-sectional estimates with follow-up data, the study aims to provide data support for the prevention and treatment objectives under the “Healthy China 2030” initiative.

## 2. Methods and Analysis

### 2.1 Study Population

The ABC Study aims to conduct a comprehensive multi-center investigation into the economic burden of AD patients and their families across various provinces and cities in mainland China. The study population not only includes patients diagnosed with AD at the dementia stage, but also prospectively incorporates individuals in the stage of amnestic mild cognitive impairment (aMCI). This group is particularly important for exploring the economic value of early intervention in the disease. The inclusion and exclusion criteria for patients are shown in Table 1.

**Table 1.** Inclusion and Exclusion Criteria of Patients.

| Inclusion criteria | Exclusion criteria |
| --- | --- |
| ● Diagnosed according to the National Institute on Aging-Alzheimer’s Association (NIA-AA) 2011 criteria for aMCI and AD.<br>● Clinical Dementia Rating (CDR) score greater than or equal to 0.5<br>● Have good visual, auditory, and language functions, or can complete neuropsychological assessments after correction.<br>● Participants or their legal representatives sign informed consent. | ● History of stroke with neurological focal signs and imaging findings consistent with cerebral small vessel disease (Modified Fazekas score $\geq 2$).<br>● Presence of mental or neurological developmental delay.<br>● Presence of other known conditions that may cause cognitive impairment.<br>● Diagnosis of a disease that prevents completion of cognitive assessments.<br>● Refusal to sign informed consent at baseline. |

### 2.2 Sampling Strategy

The ABC Study adopted a two-stage probability sampling design to ensure data representativeness at the national level. The first stage used medical institutions as sampling units. Considering the professionalism of AD diagnosis and treatment, the study relied on 602 officially certified cognitive centers distributed across 31 provinces, autonomous regions, and municipalities as the sampling frame [8]. This centre-based sampling not only ensured the reliability of diagnosis but also covered the main distribution areas of China’s AD medical resources.

Since Guangxi, Qinghai, and Ningxia each have only one cognitive center qualified for AD diagnosis, these hospitals are directly included in the study through certainty selection.

Xizang currently lacks hospitals with standardized diagnostic capabilities for AD. We recruited and trained locally qualified neurologists to conduct patient enrollment and data collection locally. This subset of samples will be separately labeled and subjected to sensitivity analysis.

In the sample size calculation, the study used the average annual medical expenses of AD patients as the core outcome indicator, employing a mean estimation method with relative precision calculation.

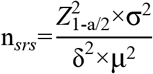

To offset the loss of independence among samples caused by Cluster Sampling, the Design Effect (DEFF) is adjusted using the intracluster correlation coefficient (rho).

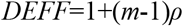

The study preset rho to be 0.005, which is a relatively robust parameter selection aimed at ensuring that the sample remains sufficiently representative even when there is a certain degree of homogeneity among hospitals. Under these assumptions, the design effect is approximately 1.50. Detailed parameters are presented in table 2.

**Table 2.** Parameters definition and selection.

| Parameters | Value |  |
| --- | --- | --- |
| Confidence level ($1-\alpha$) | 95% | According to the standard normal distribution quantile. |
| Relative error ($\delta$) | 3% | Lower than 3% |
| Population mean ($\mu$) of medical costs | 20893.39 | According to the relevant research on average economic burden of AD patients in China. |
| Population standard deviation ($\sigma$) of medical costs | 25463.5 | Reflecting the degree of dispersion of medical costs among AD patients, derived from the same referenced literature [8]. |
| Intraclass correlation coefficient ($\rho$) | 0.005 | Measures the homogeneity of medical costs among AD patients within the same hospital (cluster). As ICC in socioeconomic research usually applies as 0.001-0.005, we took a higher value to make the population more representative. |
| Average cluster size ($m$) | 100 | We applied a small-scale pre-investigation among hospitals to arrive at this average cluster size. Cluster size in each hospital is allowed to fluctuate between 40-120. The estimated average number of patients enrolled per center during the survey period is approximately 100. |

Based on the *China Alzheimer’s Disease Report 2024* and the *China Statistical Yearbook 2025* [14], we estimate the number of AD patients in each administrative region, and calculate the regional distribution proportion accordingly. The number of involving hospitals in each region is allocated proportionally according to the patient scale. The largest remainder method is used to balance the deviation caused by the integer rounding to calculate the regional hospital numbers.

The final numbers of hospitals selected in each region are shown in Table 3.

**Table 3.** Numbers of hospitals across regions.

| Region | Proportion of AD cases (%) | Hospital numbers | Patient numbers |
| --- | --- | --- | --- |
| North China | 11.681 | 11 | 1110 |
| North East China | 8.072 | 8 | 768 |
| East China | 31.801 | 30 | 3024 |
| Central South China | 27.250 | 26 | 2592 |
| South West China | 15.207 | 15 | 1446 |
| North West China | 5.989 | 6 | 570 |
| *Total* | *100.000* | *96 (rounded)* | *9510* |

*The numbers of hospitals in each region may have slight deviations due to rounding of integers. During study implementation, adjustments may be permitted without changing the regional stratification structure to ensure the overall sample size meets statistical requirements.*

### 2.3 Follow-up

To accurately assess the indirect economic costs associated with AD, a Phase 2 follow-up survey is conducted following the completion of the Phase 1 cross-sectional survey. This involves a 12-month follow-up of patients and their informal caregivers.

As caregiving time and productivity loss are cumulative and cannot be accurately captured through a single cross-sectional assessment, a nested follow-up subsample design is adopted. Based on the total Phase 1 sample size (N = 9510), approximately 20% of participant-caregiver dyads (approximately N=1902) will be selected for follow-up. A stratified random sampling strategy will be applied within clusters, ensuring that the follow-up sample maintains representativeness across regions and institutions.

Data on caregiving hours, labor participation, and productivity loss will be collected to estimate the indirect economic burden of AD. During the analysis phase, potential bias arising from selective follow-up will be addressed using weighting methods or sensitivity analyses, based on differences in key demographic and clinical characteristics between the follow-up subsample and the full Phase 1 cohort.

### 2.4. Study Variables

The ABC Study includes a multidimensional variable framework covering six domains, as shown in Table 4.

**Table 4.**
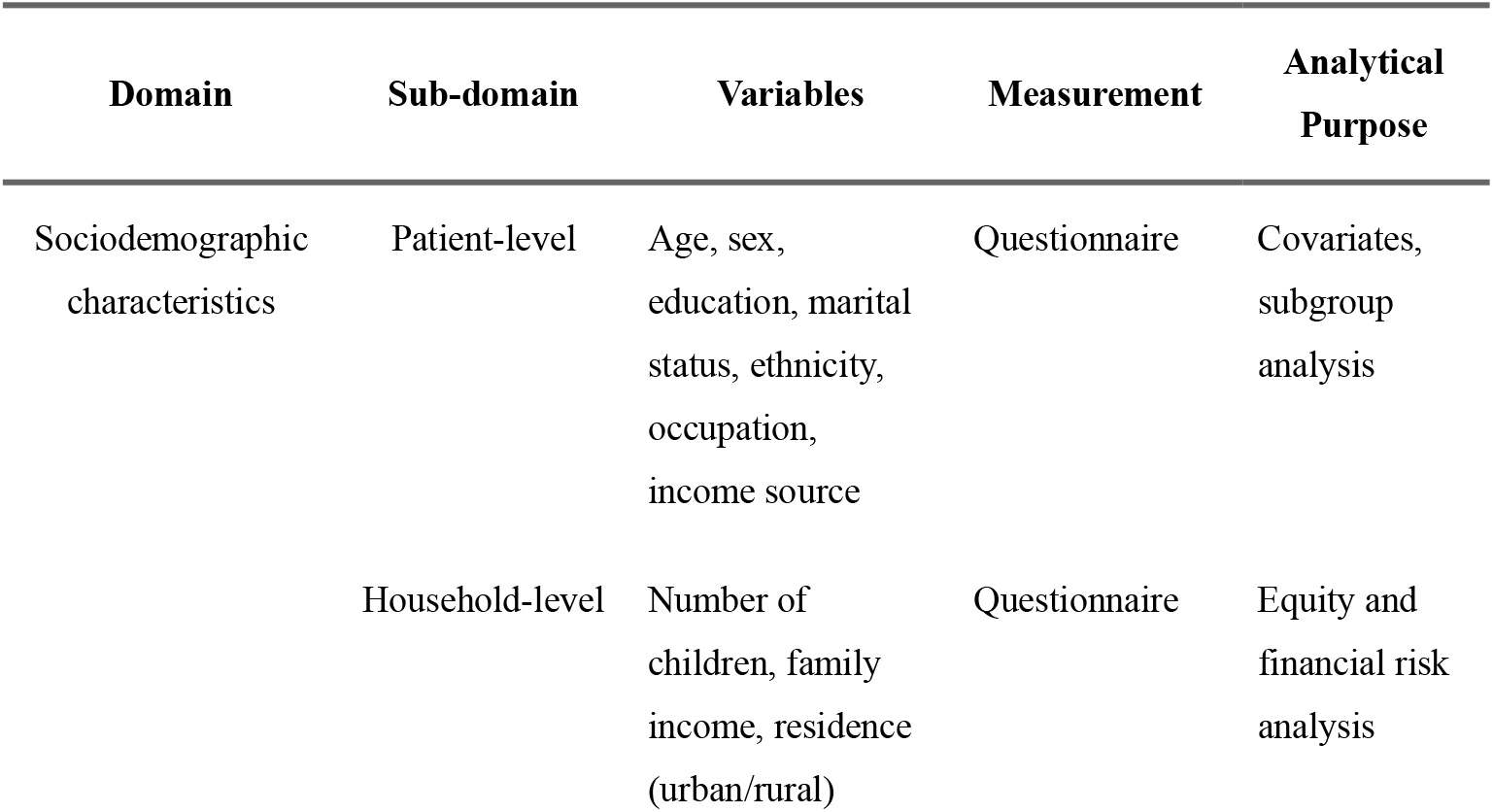
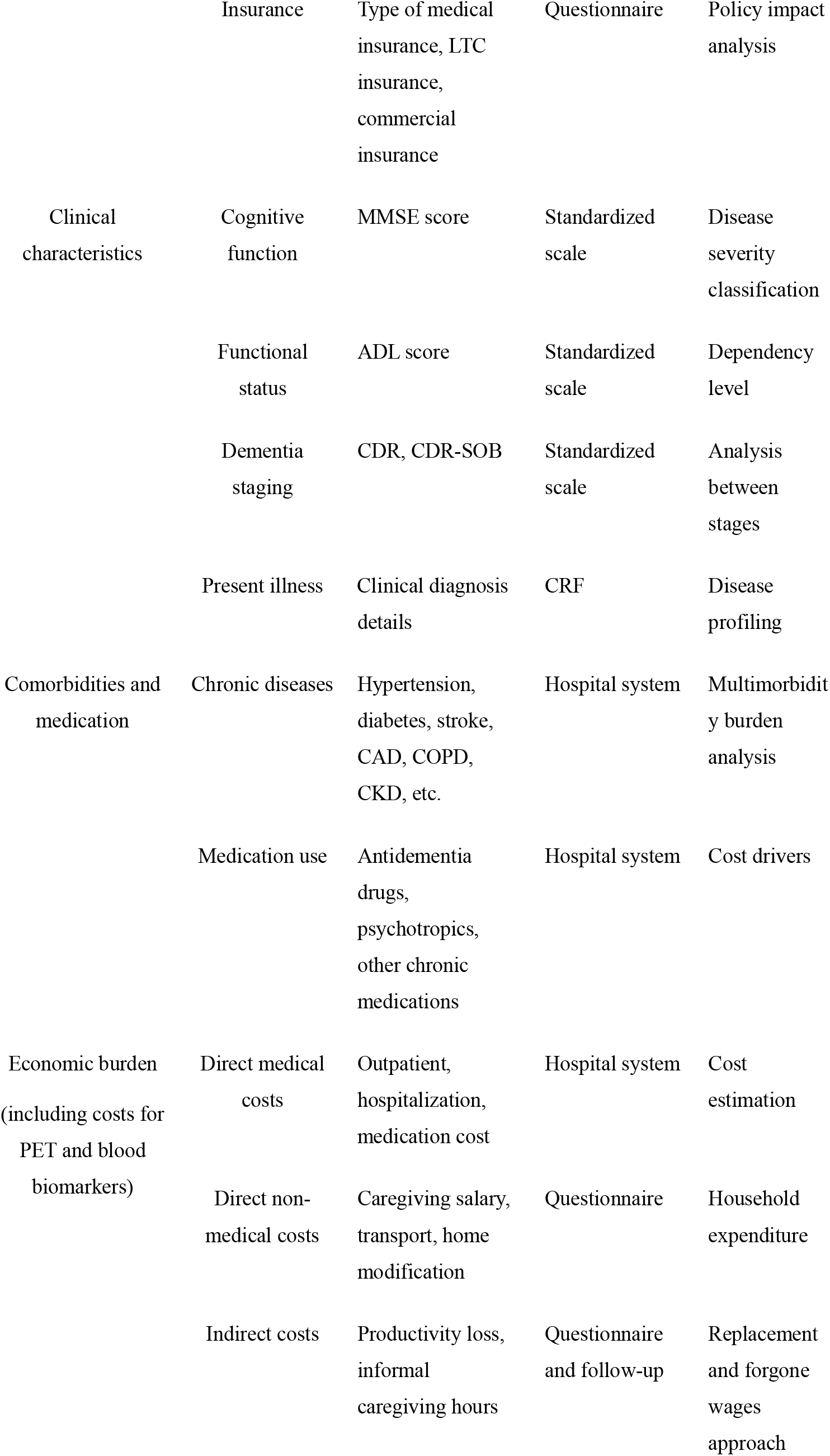
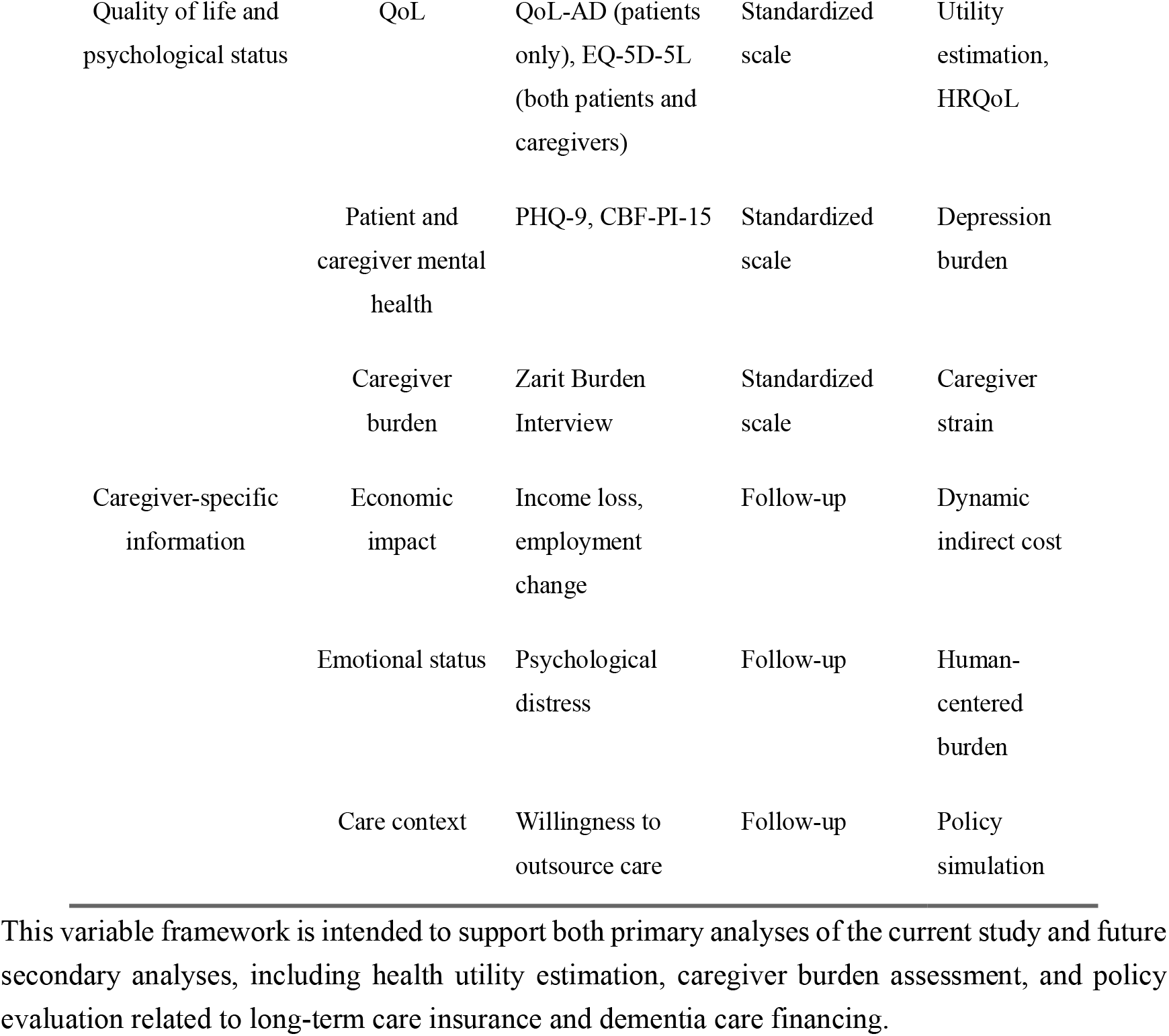
Variable framework and analytical structure.

### 2.5 Data Analysis Plan

The baseline characteristics of the study population will be described as mean ± standard deviation or median (IQR) for continuous variables and number (percentage) for categorical variables. T-test or non-parametric test will be used, and the chi-square test will be performed for comparing. Continuous cost variables will be analyzed using generalized linear models to account for potential skewness. Regression models will incorporate sampling weights, stratification, and clustering within primary sampling units (hospitals) using appropriate survey modules (e.g., Generalized Estimating Equations [GEE]).

These baseline characteristics will include age, sex, education, marital status, geographic region, place of residence (urban/rural), number of children, time of AD diagnosis, MMSE scores, ADL scores, sleep status, and the number of co-morbidities. Additional descriptive analyses will also be conducted for caregiver-level data collected in phase 2.

Missing data patterns will be examined. For variables with more than 5% missingness, multiple imputation (MI) by chained equations will be considered. Otherwise, complete-case analysis will be performed.

In the second phase, indirect cost data are collected from a subset of participants who completed follow-up interviews. To account for potential selection bias due to non-random attrition, inverse probability weighting (IPW) will be applied [11].

A logistic regression model will be used to estimate the probability of completing the second-phase survey conditional on baseline characteristics. Covariates included region, urban/rural residence, age, sex, education, marital status, household income, disease severity (CDR stage), functional status (ADL score), comorbidity burden, caregiver availability, unmet care needs, baseline direct medical and non-medical costs, and hospitalization status.

Stabilized inverse probability weights will be calculated as the marginal probability of follow-up divided by the predicted probability of follow-up. These weights will be applied to the second-phase sample to obtain estimates representative of the baseline population. Extreme weights will be examined and truncated in sensitivity analyses to assess robustness.

#### 2.5.1 Economic Burden Estimation

The per capita cost will be calculated by summing up the costs across the mentioned categories. Nationwide costs will be calculated using provincial numbers of AD patients as weights. Subgroup analysis will be used, stratified by severity (aMCI, mild, moderate, severe), age (40-59, 60-69, 70-79, ≥80). Caregiving time (hours) that absent from work will be collected by caregiving items. When estimating the economic impact on informal caregivers, two mostly used approaches (replacement approach and forgone wages approach) will be used [11], [12]. The replacement approach model is as follows:

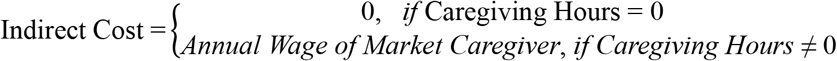

Unlike in other countries, formal care professionals in China are typically hired on a full-time, monthly salary basis. Therefore, the conventional “time × wage” approach does not fully capture the true nature of replacement costs. Based on this, we adjust our model as described above. Annual Wage of Market Caregiver is based on the 2024 average wage for the “Resident Services, Repair, and Other Services” sector in each province [13]. This standard is used to adjudicate nursing costs [11],[14]. The forgone wages approach model is as follows:

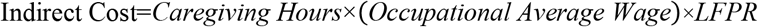

Occupational average wage will be investigated by this study, and labor force participation rate (LFPR) will be estimated using data from National Statistic Bureau [15].

Nationwide grand total costs will be calculated using national costs per capita plus the number of AD patients, and projection will be made based on the data of population structure using world population prospect 2024 by the United Nation, presuming an unchanged prevalence according to other research, or utilizing an estimated future prevalence [12].

As the study population was recruited from clinics and hospitals with diagnostic capacity, the observed sample mainly represents patients who have already accessed medical care. However, a substantial proportion of individuals with AD in China remain undiagnosed or untreated in the community. Direct extrapolation of observed per-capita costs to the national AD population may therefore overestimate direct medical costs. To improve the national representativeness of cost estimates, a post-stratification cost adjustment model will be applied to account for undiagnosed and untreated patients [17]. A conventional form of this adjustment model is expressed as:

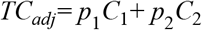

Because nationally representative cost data for undiagnosed patients with AD are not currently available in China, this study will use the ratio of the cost of undiagnosed patients to that of diagnosed patients derived from a study in Hong Kong, which may better reflect the real-world situation in Chinese populations [18]. In addition, a crude diagnostic ratio will be estimated using the one-year consultation rate reported in the China Alzheimer Report 2025 and post-consultation diagnostic accuracy reported in previous studies [19,20]. Based on published evidence, the one-year consultation rate was 32.6%, and post-consultation diagnostic accuracy was assumed to range from 61% to 73%; the mean value of 67% will be used in the main analysis. Sensitivity analyses will be conducted under alternative assumptions regarding post-consultation diagnostic accuracy and the ratio of the cost of undiagnosed patients to that of diagnosed patients. The crude diagnostic ratio will be calculated as follows:

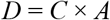

In which D denotes the crude diagnostic ratio, C denotes the one-year consultation rate, and A denotes post-consultation diagnostic accuracy. The adjusted total cost will be calculated as:

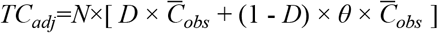

In which N denotes the total number of patients with AD, D denotes the crude diagnostic ratio, 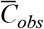 denotes the mean per-capita cost among observed diagnosed or treated patients, and θ denotes the cost ratio of undiagnosed patients relative to diagnosed patients.

#### 2.5.2 Unmet Value of Market Care

Caregiving status (needing care and being cared, needing care and not being cared, not needing care) will be collected in patients. Regional caregiving gap (percentage of unmet caregiving) will be calculated based on investigation with provincial prevalence as weight. By multiplying caregiving gap with regional number of patients and regional indirect costs estimated by approaches in 2.5.1, Regional and national unmet value of market care will be calculated. Projection will be made based on the data of population structure using the United Nations World Population Prospects 2024, presuming an unchanged prevalence according to other research, or utilizing an estimated future prevalence [5,21].

#### 2.5.3 Alzheimer’s Disease Dependency Ratio (ADDR)

Following the extension from the conventional dependency ratio (DR) and the old-age dependency ratio (OADR), the ABC Study explores the ADDR as a disease-specific indicator of potential care and economic pressure associated with AD [22,23]. The ADDR will be calculated as follows:

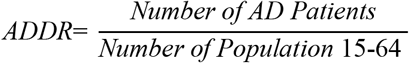

The numerator will be the estimated and projected number of individuals living with AD, as described in section 2.5.1, and the denominator will be the provincial or regional population aged 15-64 years. Provincial and regional population estimates and projections will be generated using a simplified cohort component technique based on the accessible data from the United Nations, National Bureau of Statistics, and Baidu Migration Bigdata Platform [13,21,24,25].

#### 2.5.4 AD-related Catastrophic Health Expenditure

This study will also utilize costs and income data to calculate AD-related catastrophic health expenditure (CHE), which is defined as AD-related medical costs exceeding 40% of household income, as follows:

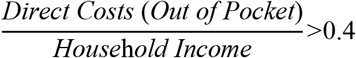

Sensitivity analysis will be performed by adjusting the percentage of CHE and replacing the household income by an estimated household capacity to pay [26].

#### 2.5.5 Quality of life and psychological status

QoL and psychological status of patients and caregivers will be analysed using validated instruments, including EQ-5D-5L, QoL-AD, PHQ-9, and the Zarit Burden Interview. Multivariate regression models will be used to identify factors related to quality of life and psychological outcomes.

#### 2.5.6 Changes in the economic burden of longitudinal follow-up of patients and caregivers

Longitudinal follow-up is designed to assess the longitudinal changes in the economic burden of patients with AD and their caregivers over time. The analysis will focus on changes in the total economic burden during disease progression, and explore the impact of disease staging and care burden on different types of cost. In addition, we will assess changes in the impact of care on families, including changes in caregiver productivity, income and care-related expenditure.

### 2.6 Patient and Public Involvement

Patients and the public are not involved in the design, or conduct, or reporting, or dissemination plans of this research.

## 3. Discussion

By constructing a comprehensive variable database covering 31 provinces, the ABC Study is designed to provide a structured evidence base for policy-making, resource allocation, and clinical pathway optimisation in dementia care in China. Compared with the data estimation of China’s AD health economics published by Jia et al. in 2018, this study improves the methodology and reflects the current situation of the health economics burden in China and the evolution of policy and social background [6,7]. The ABC Study adopts a national multi-center cluster sampling design, and plans to collect data covering all provinces in the country through standardized data collection to improve the representativeness and comparability of research samples. In addition, it incorporates a more comprehensive cost framework system, covering direct medical costs, non-medical costs and indirect costs, and includes caregiver-related outcome indicators and quality of life assessment indicators in detail. Unlike previous studies that focused mainly on the elderly population, this study is designed to include a group of young AD patients aged 45-60, some of whom had not yet retired, thereby enabling a more comprehensive assessment of productivity loss and early disease burden.

A further strength of this study is that it is the first nationwide disease burden study in China to include individuals with aMCI, this may help fill an evidence gap in early-stage AD and provide data for future HTA of early screening and interventions. In addition, previous studies often estimated replacement costs using hourly wage-based assumptions, which may not be appropriate in Chinese caregiving market, where caregivers are more commonly employed on a full-time or monthly-salary basis rather than an hourly basis[6,27]. This consideration has been incorporated into the questionnaire design, and the replacement cost approach and the forgone wages approach are designed to calculate the indirect cost.

Building on previous research of AD burden in China, the ABC Study may also provide a structured variable framework for future interdisciplinary studies, spanning health economics, caregiver research, dementia policy, and long-term care financing. The potential value of the ABC Study data is mainly reflected in the following strategic research directions.

### Future Research on Caregiver Burden and Family Resilience

Future research could use the Zarit scale and family income structure data to construct a multiple regression model for caregiver burden [28]. By analyzing the nonlinear relationship between caregiver psychological stress and family financial risk across different disease stages, families at the highest risk of burden can be identified.

Furthermore, based on 12-month follow-up data, it is possible to investigate whether caregivers’ psychological states may influence patients’ cognition through changes in care quality.

### Health Technology Assessment (HTA) and Medical Insurance Access Analysis

With the approval and launch of new biomarker-guided drugs such as Aβ monoclonal antibodies in China, healthcare insurance decision-makers are facing financial pressure and challenges in access decisions [29]. The medical expenses at different stages provided by the ABC Study are essential parameters for conducting pharmacoeconomic models.

### Policy Optimization and Expansion of LTCI

Currently, China’s LTCI pilots mainly focus on physical disability and have insufficient coverage for disability caused by cognitive impairment[30]. The ABC Study records the impairment of patients’ daily lives and the corresponding care time. Future studies can use this data to evaluate the differences in the effectiveness of different payment ratios and assessment tools in reducing family burdens by comparing the LTCI coverage models of different provinces [31].

### Catastrophic Health Expenditure (CHE) and the Risk of Poverty Due to Illness

As a long-term chronic disease, AD imposes a cumulative economic burden on families. Using the detailed household income and expenditure structure variables, the incidence of CHE among AD families in different provinces can be accurately calculated [32].

### ADDR as Indicator of Future AD-related Care Pressure

ADDR may represent a useful extension of dependency-based indicators from general demographic ageing to disease-specific burden. The conventional dependency ratio was originally developed to reflect the pressure imposed by dependent populations on the working-age population, and was later extended to old-age dependency measures in response to population ageing [22,23]. Building on this, the ABC Study further focuses on AD-related burden through the ADDR. This may be particularly critical in China, where rapid population ageing and regional migration are reshaping the balance between care needs of AD patients and the capacity of the working-age population to provide support.

## Supporting information

Appendix-Study Data Collection Forms

## Data Availability

Study data are collected using an electronic data capture system and will be available from the corresponding author upon reasonable request, subject to ethical and institutional requirements.

## Ethics and Dissemination

The original protocol received approval from the Ethics Committee of Xuanwu Hospital, Capital Medical University, Beijing, China. The study protocol has been registered at ClinicalTrials.gov and the Chinese Clinical Trial Registry(ChiCTR).

All subjects or their legal representatives signed written informed consent. For subjects with impaired cognitive function, MMSE is used by trained physicians to evaluate decision-making ability, and informed consent is signed by their legally authorised representatives on behalf of those without decision-making ability.

## Author Statement

All authors made a substantial contribution to the development of the project. Y.X.W. and M.Z.Y. wrote the manuscript. All other authors edited the manuscript.

## Acknowledgements

We would like to express our sincere thanks to all medical workers, patients and caregivers participating in this study.

