## Appendix-Study Data Collection Forms for "The Alzheimer’s disease Burden in China (ABC) study: protocol for a nationwide multicentre cross-sectional and prospective cohort study"

### Study Data Collection Forms for the ABC study

| | |
|---|---|
| Document purpose | Prepared for use as supplementary appendix material in a protocol manuscript |
| Scope of this document | Self-designed content only; standardized instruments used in the study are acknowledged below but are not reproduced in full in this appendix |
| Source materials | The main relevant issues of baseline questionnaire and second-phase follow-up questionnaire are presented below |
| Language note | Translated and lightly standardized for protocol-style English while preserving item meaning |

*Note: Standardized scales administered in the study include QoL-AD, EQ-5D-5L, PHQ-9, CBF-PI-15, MMSE, ADL, CDR, and the Zarit Burden Interview. These scales are part of study data collection; however, their full item content is not presented in this document.*

*QoL-AD, MMSE, ADL and CDR are administered to patients only; Zarit Burden Interview is administered to caregivers only; EQ-5D-5L, PHQ-9, CBF-PI-15 are administered to both patients and caregivers.*

### Appendix A. Baseline Case Report Form and Core Questionnaire

This appendix contains the translated non-scale content from the baseline case report form and related self-designed questionnaire modules.

#### 1. Opening Statement / Introductory Script

- Dear participant and family member, we are conducting a nationwide survey on the burden of Alzheimer's disease (AD) in China. The purpose of this study is to understand the economic and caregiving burden faced by people living with AD and their families, so as to provide useful evidence for future treatment, prevention, and health policy development related to AD.
- This study has been approved by the Ethics Committee of Xuanwu Hospital, Capital Medical University. We will strictly comply with all relevant laws, regulations, and ethical requirements. This questionnaire is for research purposes only. Your personal information and responses will be kept strictly confidential and will not be disclosed or used for any other purpose. You have the right to withdraw from the survey at any time without providing any reason.
- We sincerely invite you to participate in this survey. Your participation will provide important information for improving AD prevention, control, and caregiving in China. If you have any questions or concerns, please feel free to contact us. Thank you for your participation.

#### 2. Study Team Contact Information

#### 3. Informed Consent Statement

- I have carefully read the above introductory statement and understand the purpose, content, procedures, and confidentiality arrangements of this survey.
- I understand that participation in this survey is entirely voluntary. If I have any questions or concerns, I may contact the study team at any time.
- I understand that my participation is important to this research, and I also understand that I may withdraw from the survey at any time without any adverse consequences.
- I agree to participate voluntarily, and I understand that my survey responses will be used for academic research purposes only and not for any other purpose.
- Participant signature / family member signature: __________
- Date: __________

#### 4. Eligibility Review

##### Inclusion Criteria

1. Diagnosed according to the National Institute on Aging–Alzheimer’s Association (NIA-AA) criteria for aMCI and AD.
2. Clinical Dementia Rating (CDR) score greater than or equal to 0.5
3. Have good visual, auditory, and language functions, or can complete neuropsychological assessments after correction.
4. Participants or their legal representatives sign informed consent.

*If any of the above items is answered "No," the participant is not eligible for inclusion in this study.*

##### Exclusion Criteria

1. History of stroke with neurological focal signs and imaging findings consistent with cerebral small vessel disease (Modified Fazekas score ≥2).
2. Presence of mental or neurological developmental delay.
3. Presence of other known conditions that may cause cognitive impairment.

4. Diagnosis of a disease that prevents completion of cognitive assessments.
5. Refusal to sign informed consent at baseline.

*If any of the above items is answered "Yes," the participant is not eligible for inclusion in this study.*

- Does the participant meet the eligibility criteria specified in the protocol? Yes / No
- Investigator signature: __________ Date: __________

##### 5. Visit Information

- Patient name
- Patient sex: Male / Female
- Place of residence (province, city, district/county)
- Area of residence: Urban / Rural
- Name of study hospital (full name)
- Name of interviewer
- Date of interview: Year / Month / Day

##### 6. Patient Basic Information

- Date of birth: Year / Month
- Education level
- Ethnicity
- Marital status
- Retirement status
- Occupation before retirement
- Annual income in the most recent year
- Number of children
- Number of household members living together
- Caregiving Status (Needing & having received care, Needing & not having received care, Not needing care)
- Number of caregivers (If having received care)

##### 7. Caregiver Information Within the Main CRF

- Relationship to the patient
- Occupation
- Average number of caregiving episodes per month
- Average duration of each caregiving episode
- Average working time occupied by each caregiving episode
- Annual income in the most recent year
- Additional expenditures due to caregiving
- Medical expenditures of the caregiver in the past year

##### 8. Patient Health Behaviors, Care Environment, and Insurance

- Smoking status
- Alcohol use
- Average daily exercise time
- Average daily sleep duration
- Nutritional status
- Awareness of digital therapeutics for cognition
- Care setting

- Insurance type

#### 9. Clinical Information

- MMSE score
- ADL score
- CDR scores, including domain scores, sum of boxes, and global CDR
- Clinical diagnosis (aMCI, mild AD, moderate AD or severe AD)
- Specific manifestations of memory impairment
- Degree to which cognitive impairment affects daily living and social functioning
- Past medical history

#### 10. Economic Burden Information

| Domain | | Items |
|---|---|---|
| Direct medical costs | Outpatient costs | Reason for outpatient visit; number of visits in the past 3 months; total cost of the most recent AD-related outpatient visit; registration fee; diagnostic / assessment fee; imaging costs (including costs of PET); laboratory test costs (including costs of blood/CSF biomarkers); medication costs; digital diagnosis / treatment costs; non-pharmacological treatment costs; out-of-hospital medication expenditure. |
| | Inpatient costs | Reason for hospitalization; number of AD-related hospitalizations in the past 12 months; expenditure for the most recent hospitalization; bed charges; nursing fees; laboratory examination fees (including costs of blood/CSF biomarkers); imaging examination fees (including costs of PET); treatment fees; medication fees; surgical fees; non-pharmacological treatment fees; out-of-hospital medication expenditure. |
| Direct non-medical costs | | Paid care costs; long-term care insurance reimbursement where applicable; health and wellness products costs; family facility modification and repair costs. |
| Indirect/ caregiver-related costs | | Transportation and meal expenses related to medical visits; caregiving time; relationship of informal caregiver to the patient; lost working hours; wage loss; caregiver mental illness or worsening of mental illness; economic loss due to accidental injury. |

#### 11. Primary Caregiver Questionnaire in the Main CRF (Self-designed Non-scale Part)

- Education level
- Marital status
- Retirement status
- Occupation or occupation before retirement
- Number of children
- Health status
- Type of chronic disease
- Health behaviors
- Whether the caregiver received caregiving training in the past year
- Whether the caregiver received psychological counseling services in the past year

### Appendix B. Second-Phase Follow-up Questionnaire

This appendix contains the translated non-scale content from the second-phase follow-up questionnaire focusing on caregivers of AD patients including work loss, caregiver time costs, household income changes, paid care, and willingness-to-pay items.

#### 1. Participant and caregiver identification

- Hospital name
- Participant ID number
- Relationship of respondent to the patient
- Is the patient retired?
- Did the patient retire normally upon reaching retirement age?
- Did the patient retire early because of AD?
- If yes, how many years early did the patient retire?
- If the patient is still working, is early retirement due to AD expected?
- If yes, in how many years is early retirement expected?
- Has the patient been transferred to another position with reduced salary because of AD?
- What was the original annual salary?
- What is the annual salary after the reduction?
- For how many years has the salary been reduced?
- Has the patient retired from formal employment but remained engaged in other work?
- When was the patient first diagnosed with AD? Please specify year and month.
- What is the relationship between the primary caregiver and the patient?
- Age of the primary caregiver
- Sex of the primary caregiver

#### 2. Caregiver time loss and income loss

- Average monthly number of outpatient or emergency visits accompanied by the primary caregiver in the past year
- Average monthly working hours missed because of accompanying the patient to outpatient or emergency visits
- Average monthly wage or income loss due to accompanying the patient to outpatient or emergency visits
- Average monthly number of hospitalizations accompanied by the primary caregiver in the past year
- Average monthly working hours missed because of accompanying the patient during hospitalization
- Average monthly wage or income loss due to accompanying the patient during hospitalization
- Average monthly working hours missed because of home-based care and companionship
- Average monthly wage or income loss due to home-based care and companionship
- Amount by which the caregiver's monthly salary decreased because of caregiving

#### 3. Care burden perception and willingness to hire support

- Are you willing to provide care related to the patient's hygiene needs?
- How do you feel when dealing with hygiene-related caregiving tasks?
- If a professional were available to take over hygiene-related caregiving tasks, would you be willing to hire such a person?
- If yes, how much would you be willing to pay per month?
- Has the caregiver sought hospital treatment for psychological problems caused by or worsened by caring for a patient with AD?
- If yes, what were the treatment expenses in the past 12 months?

##### 4. Household income and care expenditures

- Total household income of co-residing family members before diagnosis
- Total household income of co-residing family members after diagnosis
- Disposable household income after essential expenditures
- Patient's clinical diagnosis
- Average monthly cost of medical visits for the patient
- Average monthly transportation cost related to medical visits
- Average monthly meal cost related to medical visits
- Average monthly accommodation cost related to medical visits
- Are paid service providers such as nannies employed?
- If yes, what is the monthly expenditure on such paid service providers?

##### 5. Care needs, paid care, and market-price items

- What are the patient's current major caregiving needs?
- Can the current informal care provided by family members meet the patient's daily needs?
- Have you previously hired a caregiver for the patient?
- If yes, what was the average monthly payment?
- If no, what were the reasons for not hiring one?
- Have you previously learned about the market price of hiring a caregiver?
- If yes, what was the average monthly market price?
- If local nursing aides could provide the same care currently provided by family members, what is the maximum monthly amount your family would be willing to pay?

##### 6. Patient psychological condition and accident-related economic loss

- Does the patient have a psychological disorder?
- If yes, has it worsened in the past 12 months?
- In the past 12 months, how much did the patient spend on treatment for the psychological disorder?
- In the past 12 months, what was the economic loss caused by accidental injury to the patient?
